# The Role of Hydroxychloroquine in the Age of COVID-19: A Periodic Systematic Review and Meta-Analysis

**DOI:** 10.1101/2020.04.14.20065276

**Authors:** Amir Shamshirian, Amirhossein Hessami, Keyvan Heydari, Reza Alizadeh-Navaei, Mohammad Ali Ebrahimzadeh, George W. Yip, Roya Ghasemian, Meghdad Sedaghat, Hananeh Baradaran, Soheil Mohammadi Yazdi, Elham Aboufazeli, Hamed Jafarpour, Ehsan Dadgostar, Behnaz Tirandazi, Keyvan Karimifar, Aida Eftekhari, Danial Shamshirian

## Abstract

**Background:** Coronavirus Disease 2019 (COVID-19) has become a major global issue with rising the number of infected individuals and mortality in recent months. Among all therapeutic approaches, arguments have raised about hydroxychloroquine (HCQ) efficacy in the treatment of COVID-19. We carried out a systematic review and meta-analysis overcome the controversies regarding the effectiveness of hydroxychloroquine in the treatment of COVID-19.

**Methods:** A systematic search was performed in PubMed, Scopus, Embase, Cochrane Library, Web of Science, Google Scholar and medRxiv pre-print database using all available MeSH terms for COVID-19 and hydroxychloroquine up to July 19, 2020. Studies focused on the effectiveness of HCQ with/without azithromycin (AZM) in confirmed COVID-19 patients were entered into the study. Two researchers have independently evaluated quality assessment of the studies and abstracted data for data extraction. Extracted data were analyzed using CMA *v*. 2.2.064. Heterogeneity was assessed using the *I*-squared (*I*^*2*^) test, and fixed/random-effects model was used when appropriate for pooling of studies.

**Results:** Out of 26 studies entered into our systematic review, 21 studies including 14 comparative studies with control group and seven observational studies containing 103,486 participants have entered into the meta-analysis. The results of the meta-analysis on comparative studies indicated no significant clinical effectiveness (negative in RT-PCR evaluation) for HCQ regimen in the treatment of COVID-19 in comparison to control group (RR: 1.03, 95% CI, 0.79-1.34). The same result was observed for the combination of HCQ+azithromycin (RR: 1.26, 95% CI, 0.91-1.74). No significant differences were found for both HCQ (RR: 0.92, 95% CI, 0.72-1.16) and HCQ+AZM (RR: 1.72, 95% CI, 0.86-3.42) mortality rate; however, mortality was affected by age differences according to meta-regression analysis (P<0.000001). No substantial difference was observed for disease exacerbation (RR: 1.23, 95% CI, 0.65-2.30) between HCQ group and controls. Also, radiological findings significantly improved in the HCQ group (OR: 0.32, 95% CI, 0.11-0.98). Odds of known HCQ adverse effects (diarrhea, vomiting, blurred vision, rash, headache, etc.) occurred in the HCQ regimen group was approximately 3.5 times of control group (OR: 3.40, 95% CI, 1.65-6.98), but no substantial differences were found regarding intubation odds between HCQ group and control group (OR: 2.11, 95% CI, 0.31-14.03). Meta-analysis indicated no significant prophylactic effects for HCQ (OR: 0.40, 95% CI, 0.04-3.65)

**Conclusion:** This systematic review and meta-analysis showed no clinical benefits regarding HCQ treatment with/without azithromycin for COVID-19 patients. Although mortality rate was not significantly different between cases and controls, frequency of adverse effects was substantially higher in HCQ regimen group. However, due to that most of the studies were non-randomized and results were not homogenous, selection bias was unavoidable and further large randomized clinical trials following comprehensive meta-analysis should be taken into account in order to achieve more reliable findings. Also, it is worth mentioning that if this work does not allow to quantify a “value” of the HCQ, it allows at least to know what is not the HCQ and that it would be prudent not to continue investing in this direction.

## Introduction

A novel coronavirus emerged from Wuhan, China, in December 2019 has named respiratory syndrome coronavirus 2 (SARS-CoV-2) and declared as a pandemic by World Health Organization (WHO) on March 26, 2020 ^1^. According to WHO Coronavirus Disease 2019 (COVID-19) Dashboard, this novel virus has been responsible for approximately 14,765,256 infections and 612,054 death worldwide up to 6:18pm CEST, 22 July 2020.

Although a few months have passed since the onset of the new challenging disease, there is still no specific preventive and therapeutic approach in this regard. Therefore, the quarantine approach, personal hygiene, and social distancing are the basic protective measures against COVID-19 according to WHO advise for the public ^2^.

Moreover, according to a large amount of ongoing research regarding this pandemic issue, many controversies are arising daily among different fields of sciences, which has confronted a ***“pandemic”*** with an ***“infodemic”*** (e.g. *Is coronavirus an airborne? Is COVID-19 transmitted vertically in pregnancy? Should everyone wear a mask? How long can the virus survive on surfaces? Is it possible to get COVID-19 for a second time?* etc.).

In this regard, one of the hottest controversies is the hydroxychloroquine (HCQ) efficacy with/without azithromycin (AZM) for COVID-19 patients. While several studies are talking about promising effects of HCQ regimen against SARS-CoV-2 infection for both prevention and treatment ^3-5^, others try to come up with neutral or even harmful effects of this drug for such patients when there is no ample evidence ^6^. It is unavoidable that all these controversies affect the patient’s outcome significantly.

Although there are some systematic reviews and meta-analysis in this regard ^7,8^, they suffer from several limitations and flaws that should be addressed with a more comprehensive study. Hence, due to the importance of the subject, we carried out a rapid systematic review and meta-analysis, which will be updated periodically to report more robust evidence, which might help to overcome the controversies about the effectiveness of HCQ against COVID-19.

## Method

### Study design & information

The current study has designed by ***The Tabari Systematic Review Group (TSRG)*** with an updating approach named ***“Periodic Systematic Review and Meta-analysis”***, which will be updated from time to time whenever a new substantial study is published.

### Search Strategy

The Preferred Reporting Items for Systematic Reviews and Meta-Analyses (PRISMA) guideline was followed for study design, search protocol, screening, and reporting. A systematic search was performed *via* databases of PubMed, Scopus, Embase, Cochrane Library, Web of Science and Google Scholar (intitle) as well as pre-print database of medRxiv up to July 19, 2020. Moreover, gray literature and references of eligible papers were considered for more available data in this case. The search strategy included all MeSH terms and free keywords found for COVID-19, SARS-CoV-2, and hydroxychloroquine. There was no time/location/ language limitation in this search.

### Criteria study selection

Two researchers (A.H and K.H) have screened and selected the papers independently and discussed to solve the disagreements with the third-party (R.A/D.Sh). Studies met the following criteria included into meta-analysis: 1) comparative or non-comparative clinical studies including observational/interventional studies with retrospective/prospective nature with/without control group as well as Randomized Clinical Trials (RCTs); and 2) studies reported the effect of HCQ with/without AZM in confirmed COVID-19 patients. Studies were excluded if they were: 1) animal studies, reviews, case reports, and *in vitro* studies; 2) duplicate publications; and 3) insufficient for calculating of desired parameters.

### Data extraction & quality assessment

Two researchers (A.Sh and A.H) have independently evaluated quality assessment of the studies and extracted data from selected papers. The supervisors (D.Sh/MA.E) resolved any disagreements in this step. Data extraction checklist included the name of the first author, publication year, region of study, number of patients, number of controls, mean age, treatment option, medication dosage, treatment duration, adverse effects, radiological results, nasopharyngeal culture status through Reverse Transcription-Polymerase Chain Reaction (RT-PCR) and mortality.

The Jadad scale, ROBINS-*I* tool and Newcastle-Ottawa Scale (NOS) checklists were used to value the selected randomized controlled trials, non-randomized controlled trials and observational studies respectively concerning various aspects of the methodology and study process. Risk-of-bias plots have been created through the *robvis* online tool ^9^.

### Targeted outcomes

1) Clinical effectiveness of HCQ with/without AZM in the treatment of COVID-19; 2) Mortality rates; 3) Disease exacerbation; 4) Frequency of known HCQ adverse effects occurred during treatment; 5) Intubation need; 6) Radiological improvement; 7) prophylactic effects of HCQ

### Comparisons

HCQ in comparison to a control group with standard treatment; 2) HCQ+AZM in comparison to control group with standard treatment.

### Definitions

- **Clinical effectiveness:** nasopharyngeal swab culture resulted negative in RT-PCR evaluation.
- **Disease exacerbation:** clinical symptoms of the disease are worsened.
- **Adverse effects:** occurrence of known symptoms related to HCQ such as diarrhea, vomiting, blurred vision, rash, headache, etc.
- **Group-A** in forest plots: the case groups who receive HCQ with/without the AZM regimen.
- **Group-B** in forest plots: the control groups without HCQ/HCQ+AZM regimen.

### Heterogeneity assessment

I-square (*I*^*2*^) statistic was used for heterogeneity evaluation. Following Cochrane Handbook for Systematic Reviews of Interventions ^10^, the *I*^*2*^ was interpreted as follows: “*0% to 40%: might not be important; 30% to 60%: may represent moderate heterogeneity; 50% to 90%: may represent substantial heterogeneity; 75% to 100%: considerable heterogeneity. The importance of the observed value of I*^*2*^ *depends on (i) magnitude and direction of effects and (ii) strength of evidence for heterogeneity (e*.*g. P-value from the chi-squared test, or a confidence interval for I*^*2*^).*”*

In case of heterogeneity, DerSimonian and Laird random-effects model was applied to pool the outcomes; otherwise, the inverse variance fixed-effect model was used. Forest plots were presented to visualize the degree of variation between studies.

### Data analysis

Statistical analysis was performed using Comprehensive Meta-Analysis (CMA) *v*. 2.2.064 software. Risk Ratio (RR) or Odds Ratio (OR) were used for outcome estimation whenever appropriate with 95% Confident Interval (CI). Fixed/random-effects model was used according to heterogeneities. In the case of zero frequency, the correction value of 0.1 was used. Meta-regression analysis was done to examine the impact of age difference on HCQ regimen group mortality RR. However, due to insufficient data we could not apply the meta-regression analysis on the other moderator variables such as sex, underlying disease, etc.

### Publication bias & sensitivity analysis

Begg’s and Egger’s tests, as well as funnel plot was used for publication bias evaluation. A *P*-value of less than 0.05 was considered as statistically significant.

According to that the sensitivity analysis known as an essential part in systematic reviews with meta-analysis to determine the robustness of the obtained outcomes to the assumptions made in the data analysis ^11^, we conducted a sensitivity analysis to examine the effect of studies that greatly influence the result, especially by their weight through excluding them from the meta-analysis.

## Results

### Study selection process

The databases search resulted in 2421 papers. Duplicated papers have been excluded, and after first step screening, 147 papers were assessed for eligibility. Finally, 31 papers entered into qualitative synthesis, of which 25 papers entered into the meta-analysis. PRISMA flow diagram for the study selection process presented in Figure 1.

**Figure 1.**
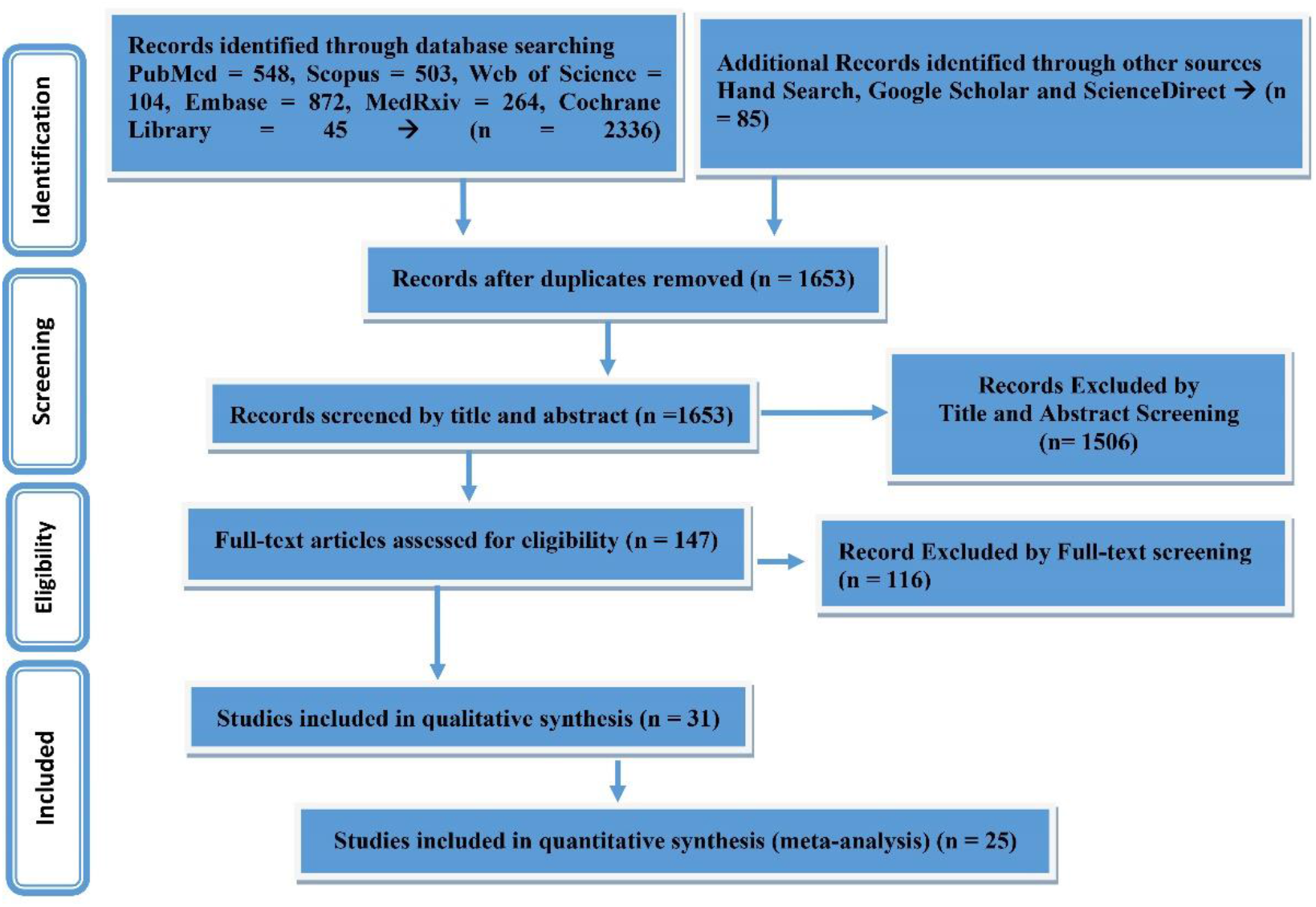
PRISMA flow diagram for the study selection process

### Study characteristics

Out of 25 studies entered into the meta-analysis, HCQ arms of the comparative studies has been combined with observational studies for effect size meta-analysis. The studies’ sample size ranged from 11 to 4716, including 24,269 participants. Characteristics of studies entered into the systematic review presented in Table 1.

**Table 1.**
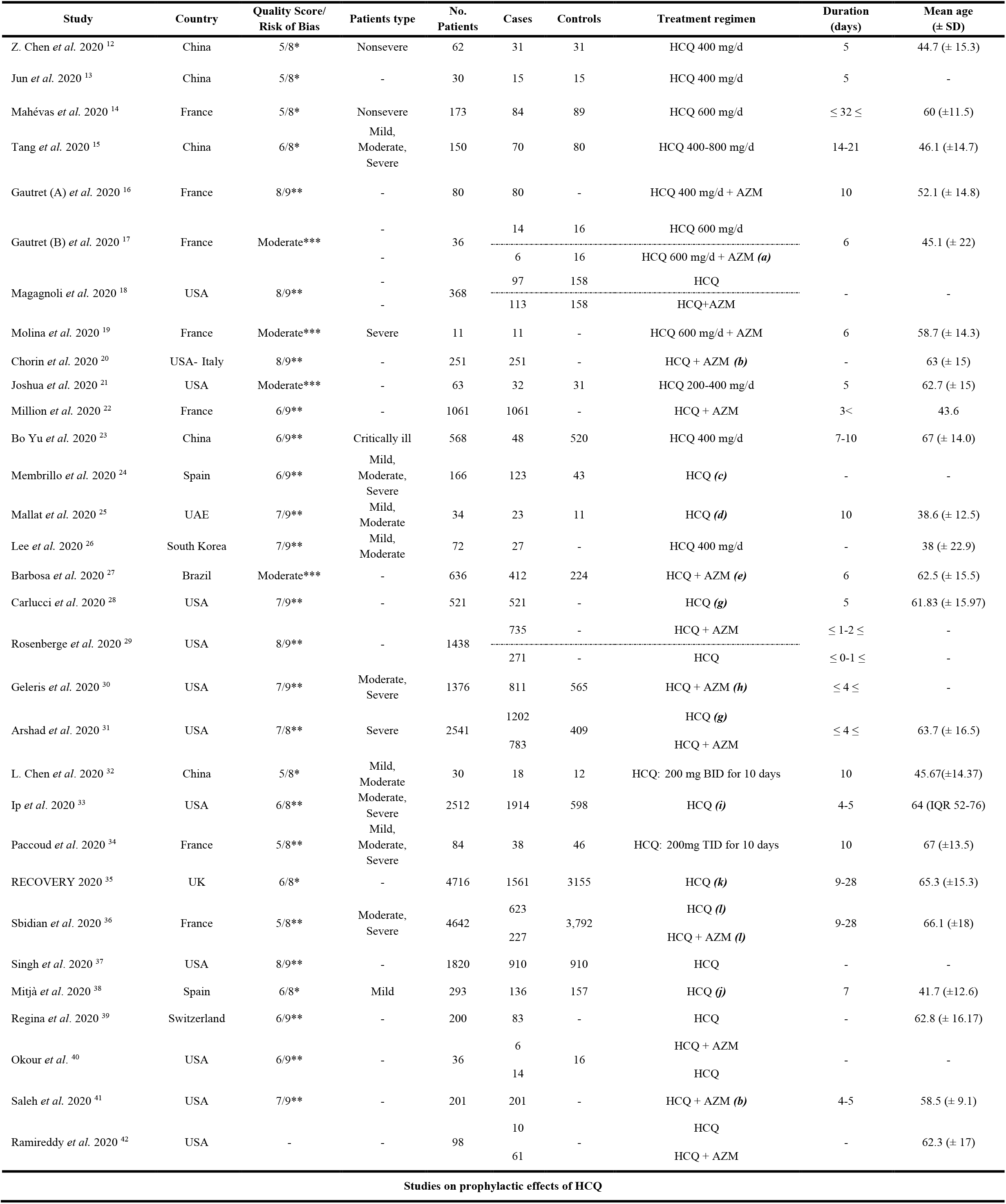
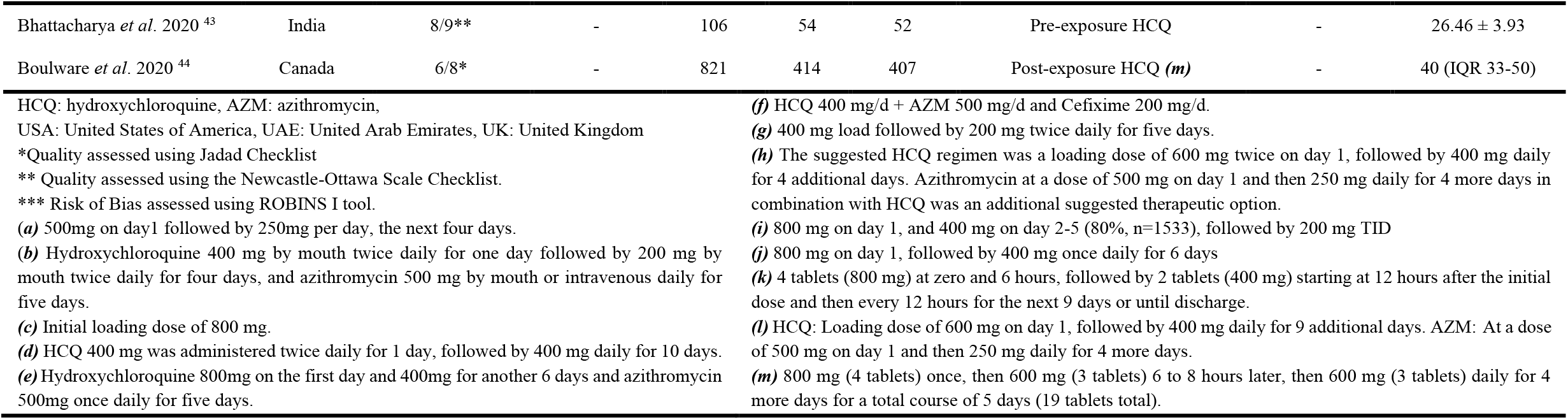
Characteristics of studies entered into the systematic review

### Quality assessment

Results of quality assessment for studies entered into meta-analysis using Jadad, ROBINS-*I* and NOS checklists were reported in Table 1 and summary of risk of bias has presented in Fig. 2.

**Figure 2.**
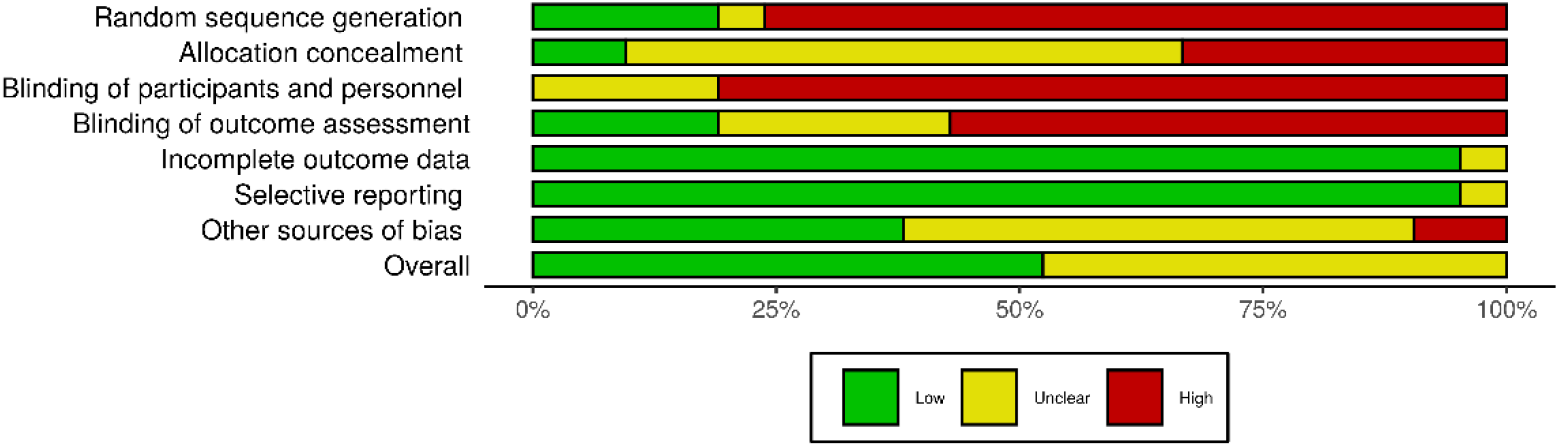
Summary of risk of bias for studies entered into the meta-analysis

### Publication bias

Results of Begg’s and Egger’s tests for every performed analysis were insignificant as follows: HCQ regimen effectiveness (*P*_*B*_=0.62; *P*_*E*_=0.34), the association between HCQ (*P*_*B*_= 0.70; *P*_*E*_=0.85) and HCQ+AZM (*P*_*B*_=0.09; *P*_*E*_= 0.43) regimen and mortality rate in controlled randomized and non-randomized studies. However, regarding overall mortality in all studies, a substantial publication bias was observed (*P*_*B*_=0.04; *P*_*E*_= 0.02). The funnel plot for publication bias of studies presented in Supplementary Fig. 1.

### Meta-analysis findings

#### Treatment outcome

##### Hydroxychloroquine regimen effectiveness

The meta-analysis of risk ratios (RR) for HCQ effectiveness in all of the comparative randomized and non-randomized studies showed that there were no significant differences between case group, who received the standard treatment with HCQ regimen and the control group that received the standard treatment without HCQ (RR: 1.03, 95% CI, 0.79-1.34). We also found no significant risk difference (RD) between two groups regarding the effectiveness of HCQ in COVID-19 patients (RD: 0.01, 95% CI, −0.14-0.17) (Fig. 3). Also, no substantial effectiveness for HCQ was found by the meta-analysis of only controlled randomized studies as well (RR: 1.19, 95% CI, 0.87-1.63; RD: 0.12, 95% CI, −0.07-0.33) (Fig. 4).

**Figure 3.**
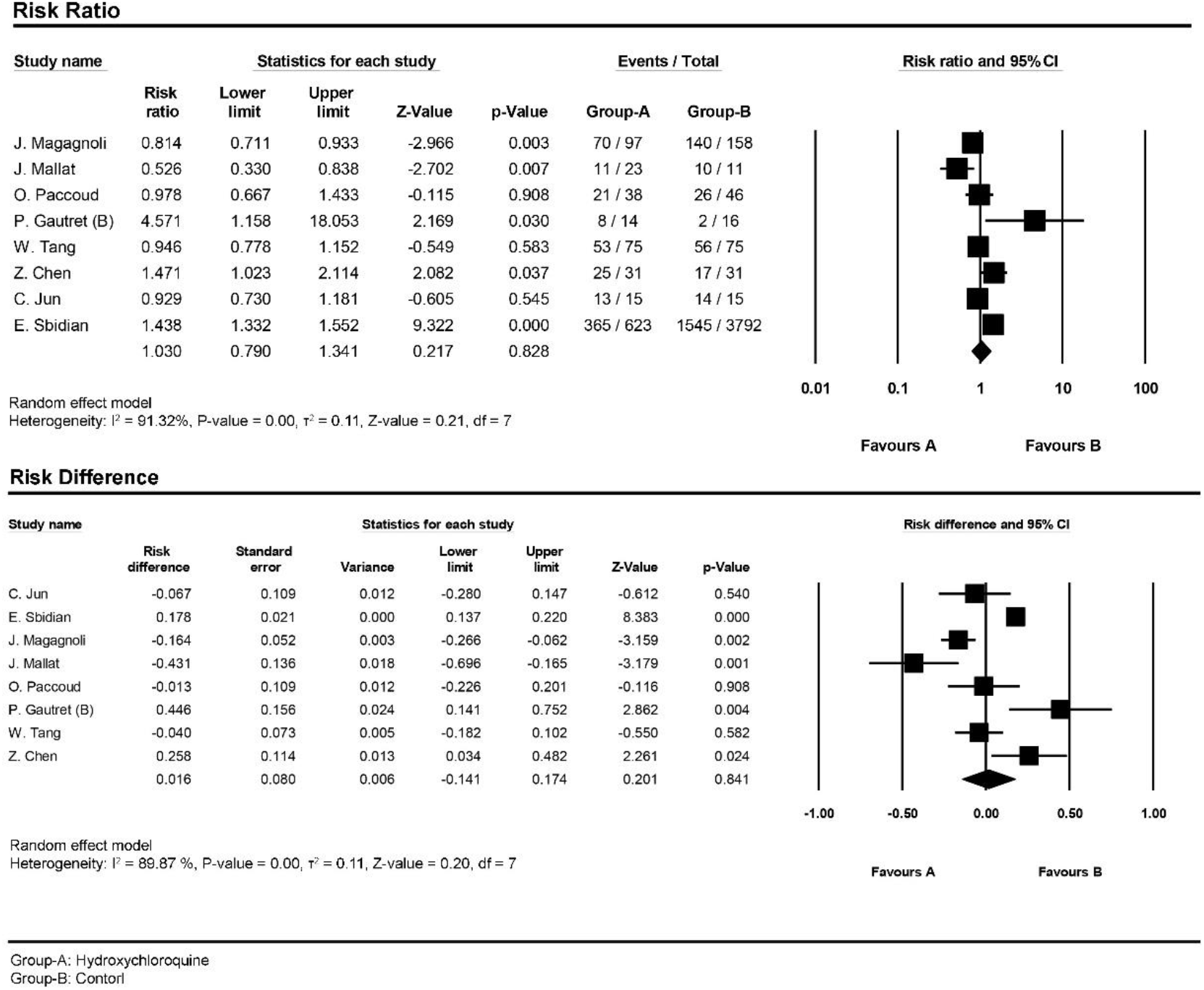
Forest plot for pooling risk ratios and risk differences regarding hydroxychloroquine regimen in comparative randomized and non-randomized studies

**Figure 4.**
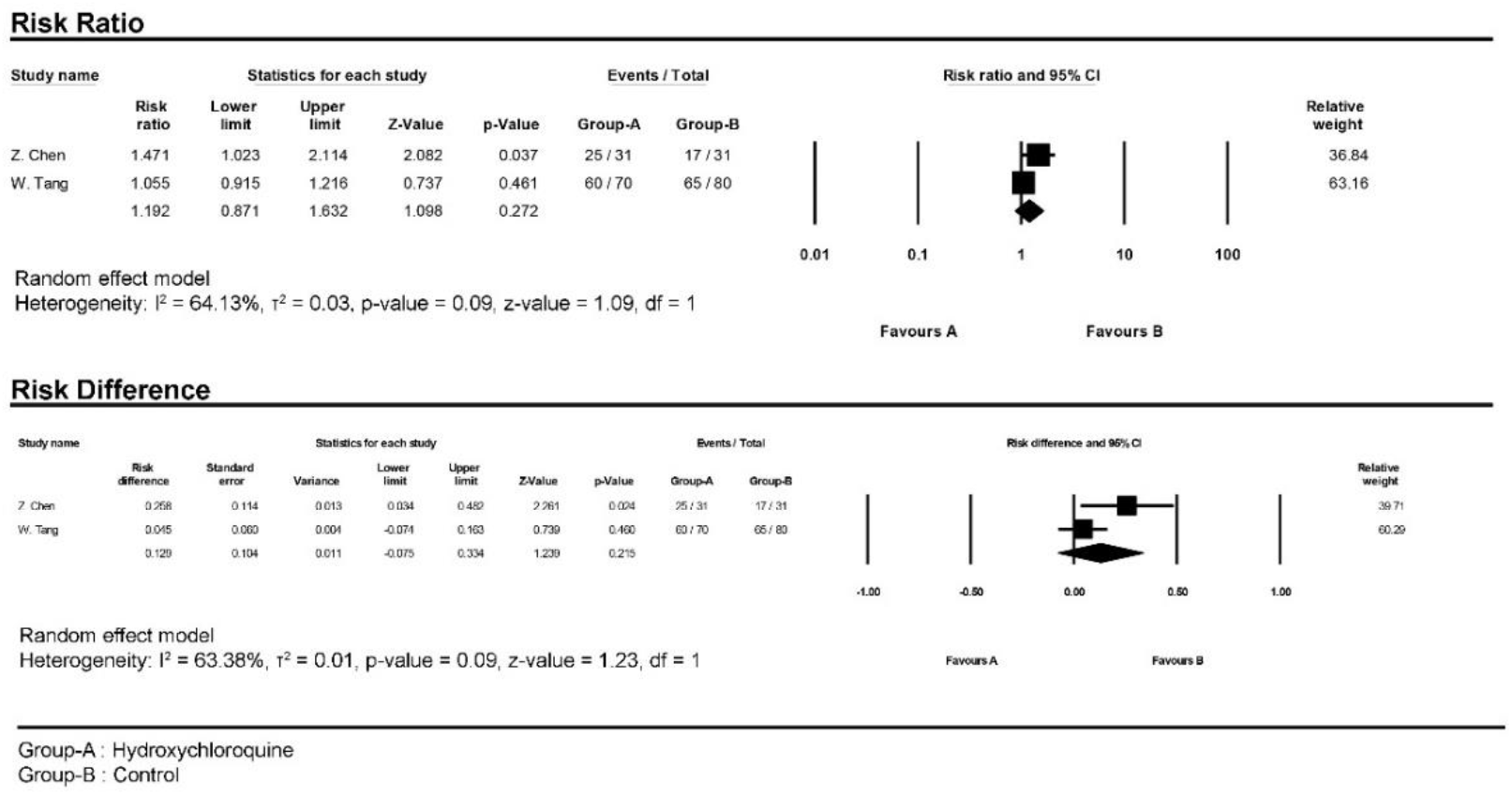
Forest plot for pooling risk ratios and risk differences regarding hydroxychloroquine regimen in only controlled randomized studies

##### Sensitivity analysis for hydroxychloroquine regimen effectiveness

Regardless of separate analysis only on controlled randomized studies (Fig. 4), to evaluate the impact of inverse RRs as well as studies’ weight on the meta-analysis results, we conducted several sensitivity analyses as follows: 1) according to the substantial relative weight of Sbidian *et al*. study the meta-analysis, by excluding this study no significant changes was observed (RR: 0.94, 95% CI, 0.77-1.15) (Supplementary Fig. 2); 2) considering five studies with *P*-value less than 0.05; three have a *P*<0.05 in favour of Group-A and two has a *P*<0.05 in favour of Group-B; these are the Magagnoli *et al*. and Mallat *et al*. studies, for which the 95% CI of RR has no intersection with the Chen *et al*., Gautret (B) *et al*. and Sbidian *et al*., thus, the new sensitivity analysis by excluding studies of Magagnoli *et al*. Mallat *et al*., resulted in no difference as well (RR: 1.17, 95% CI, 0.91-1.51) (Supplementary Fig. 3); and 4) by excluding studies in favour of Group-A, surprisingly, meta-analysis showed a significant difference (RR: 0.85, 95% CI, 0.77-0.94) (Supplementary Fig. 4).

##### Hydroxychloroquine + azithromycin regimen

No significant difference was found for effectiveness of HCQ+AZM combination regimen in comparison to control group in comparative studies entered into meta-analysis (RR: 1.26, 95% CI, 0.91-1.74); meanwhile, risk differences between groups was considerable (RD: 0.28, 95% CI, 0.01-0.54) (Fig. 5).

**Figure 5.**
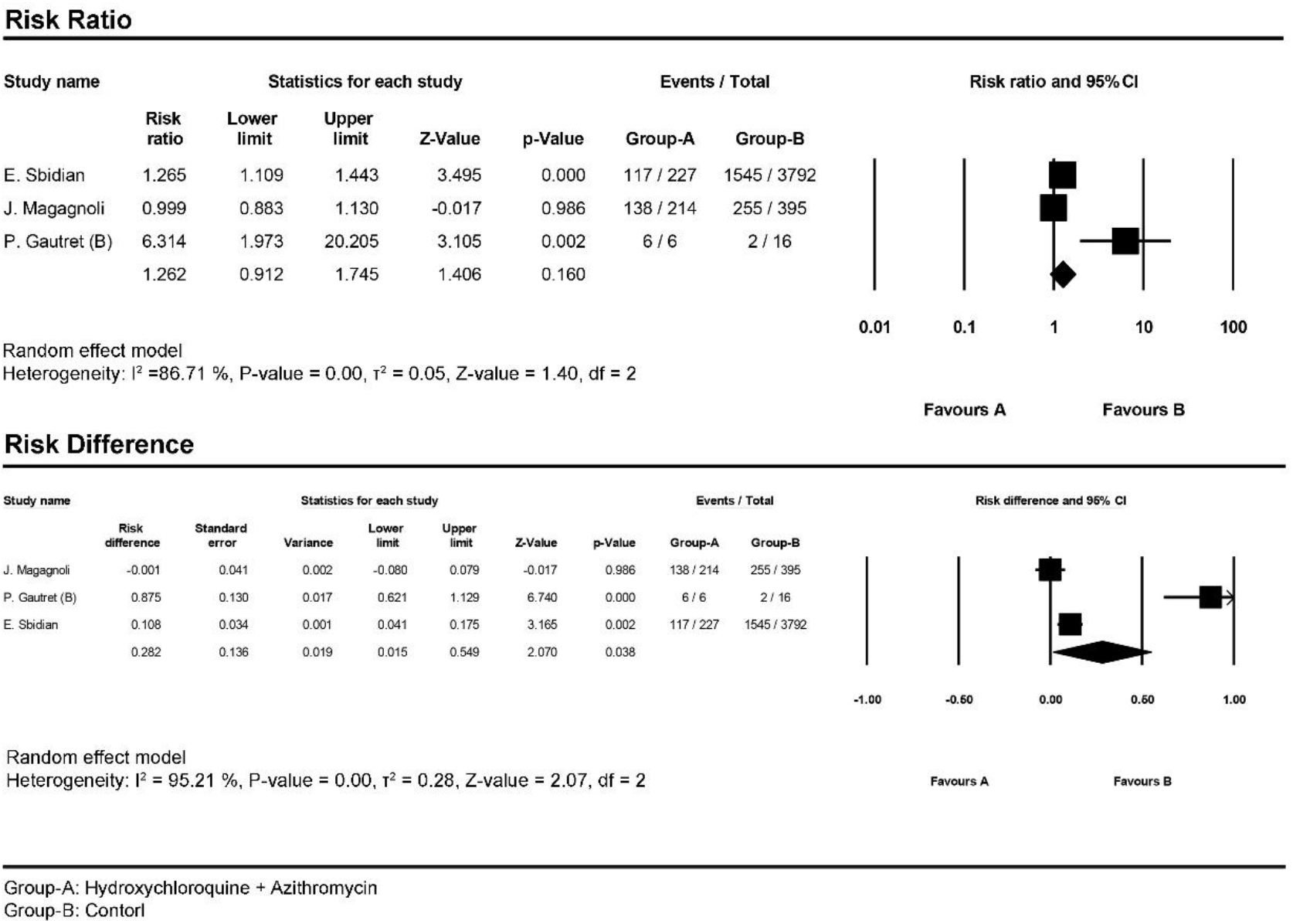
Forest plot for pooling risk ratios and risk differences regarding hydroxychloroquine + azithromycin regimen

##### Hydroxychloroquine regimen & mortality rate

The meta-analysis of death outcomes in comparative randomized and non-randomized studies showed no significant differences for mortality rate between the HCQ regimen group and standard treatment group (RR: 0.92, 95% CI, 0.72-1.16; RD: −0.01, 95% CI, −0.05-0.02) (Fig. 6).

**Figure 6.**
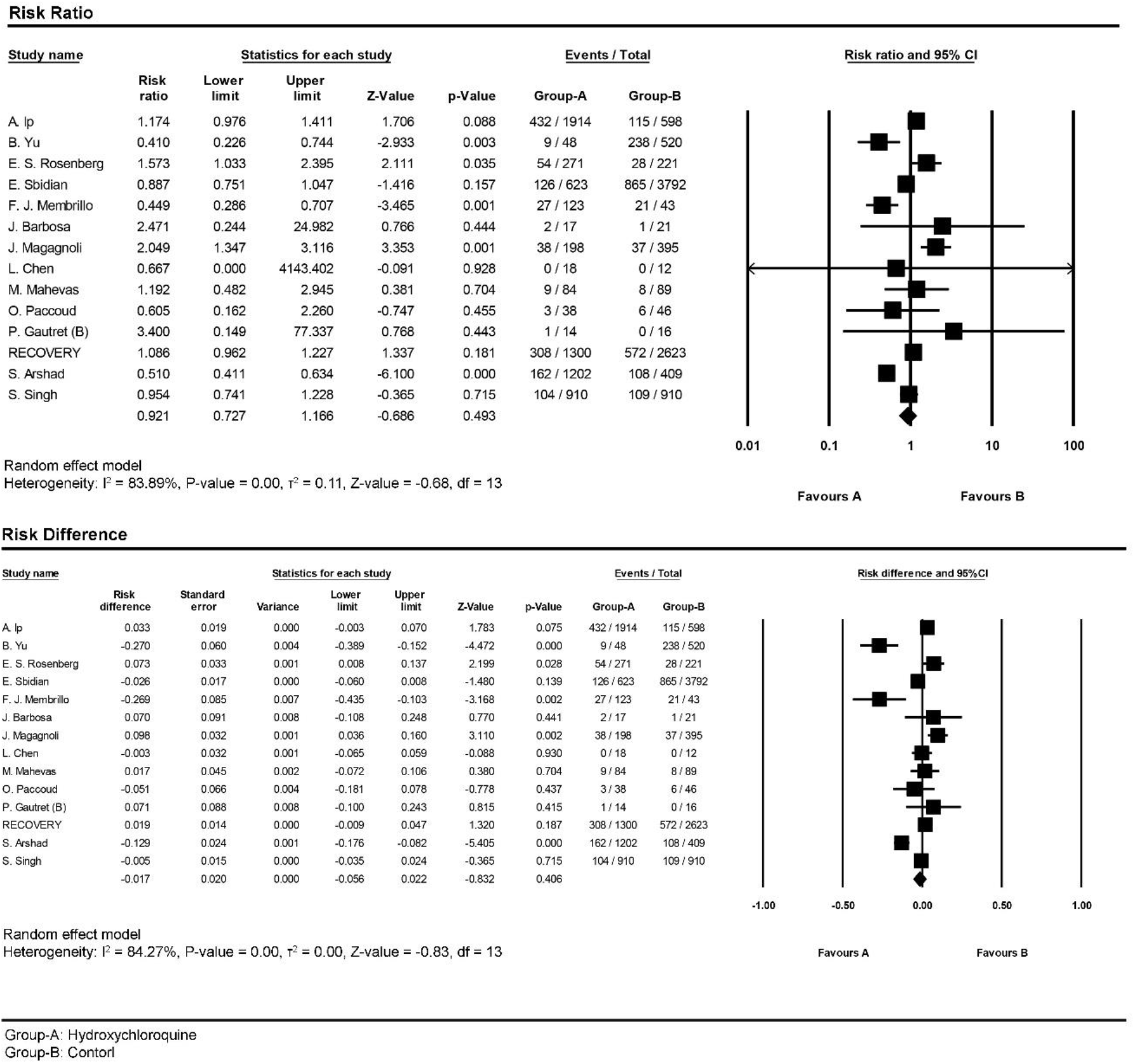
Forest plot for pooling risk ratios and risk differences regarding mortality rate in randomized and non-randomized studies

##### Sensitivity analysis for hydroxychloroquine regimen mortality

Due to the surprising mortality rate in the control group of the Yu *et al*., Membrillo *et al*. and Arshad *et al*. studies, which led to the significant inverse RR in comparison to other studies and high weight in final estimation in our analysis, we have conducted a sensitivity analysis by excluding these studies. The sensitivity analysis resulted in no significant mortality rate in the HCQ regimen arm in comparison to the control group (RR: 1.13, 95% CI, 0.97-1.32) (Supplementary Fig. 5).

##### Meta-regression analysis on the effects of age differences on mortality

Meta-regression findings indicated that age differences in studies had substantial effects on risk ratios related to the HCQ regimen group mortality rate (*P*<0.000001) (Fig. 7).

**Figure 7.**
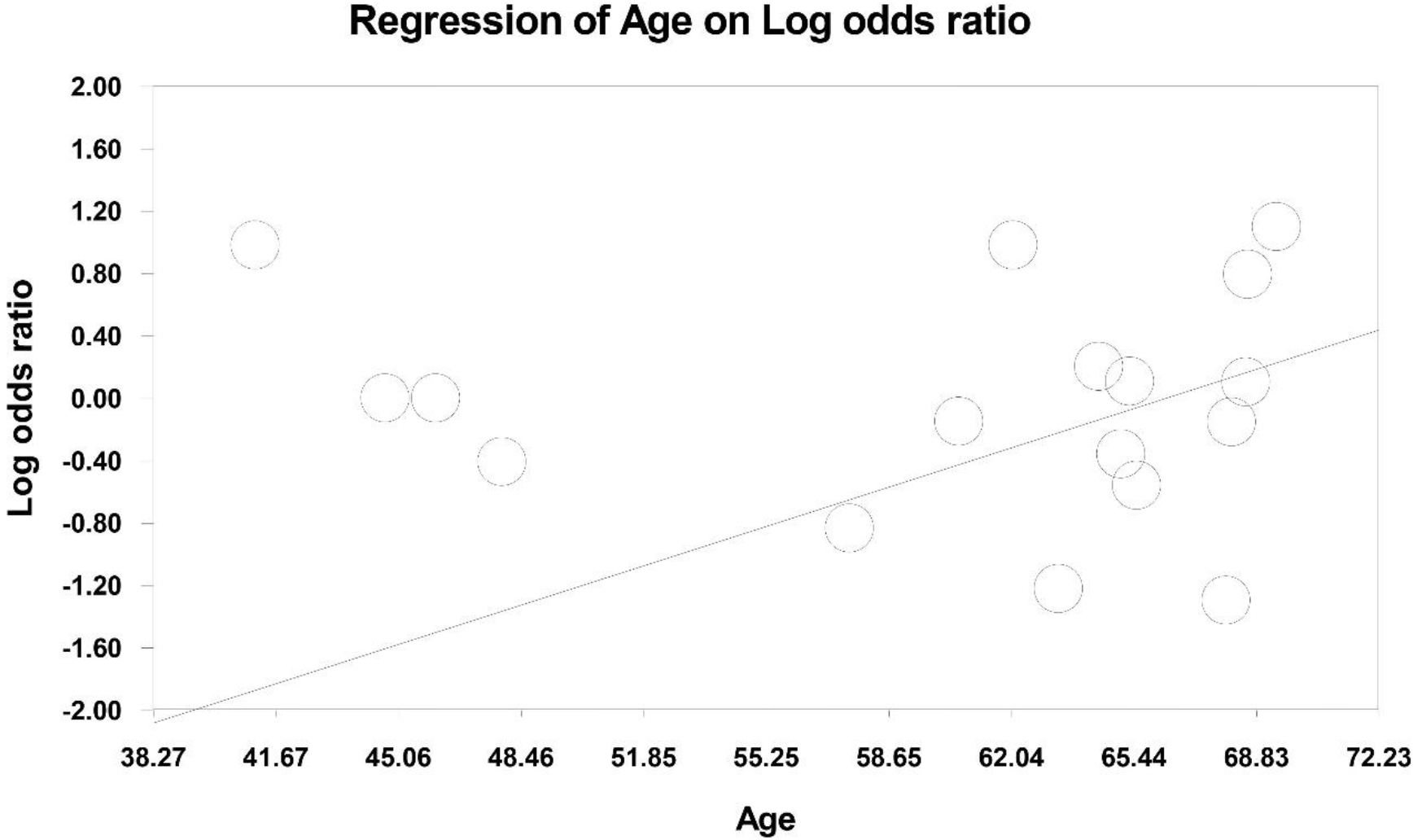
Meta-regression plot for effect of age difference on hydroxychloroquine regimen mortality Risk Ratio

##### Hydroxychloroquine + azithromycin regimen & mortality rate

The meta-analysis of mortality outcome in comparative randomized and non-randomized studies indicated no significant mortality rate in the HCQ+AZM regimen group in comparison to the control group (RR: 1.72, 95% CI, 0.86-3.42; RD: 0.13, 95% CI, −0.06-0.34) (Fig. 8). In addition, concerning substantial reverse RR of Arshad *et al*., the sensitivity analysis result by excluding the study was not significant after excluding the study (RR: 2.12, 95% CI, 0.96-4.69) (Supplementary Fig. 6).

**Figure 8.**
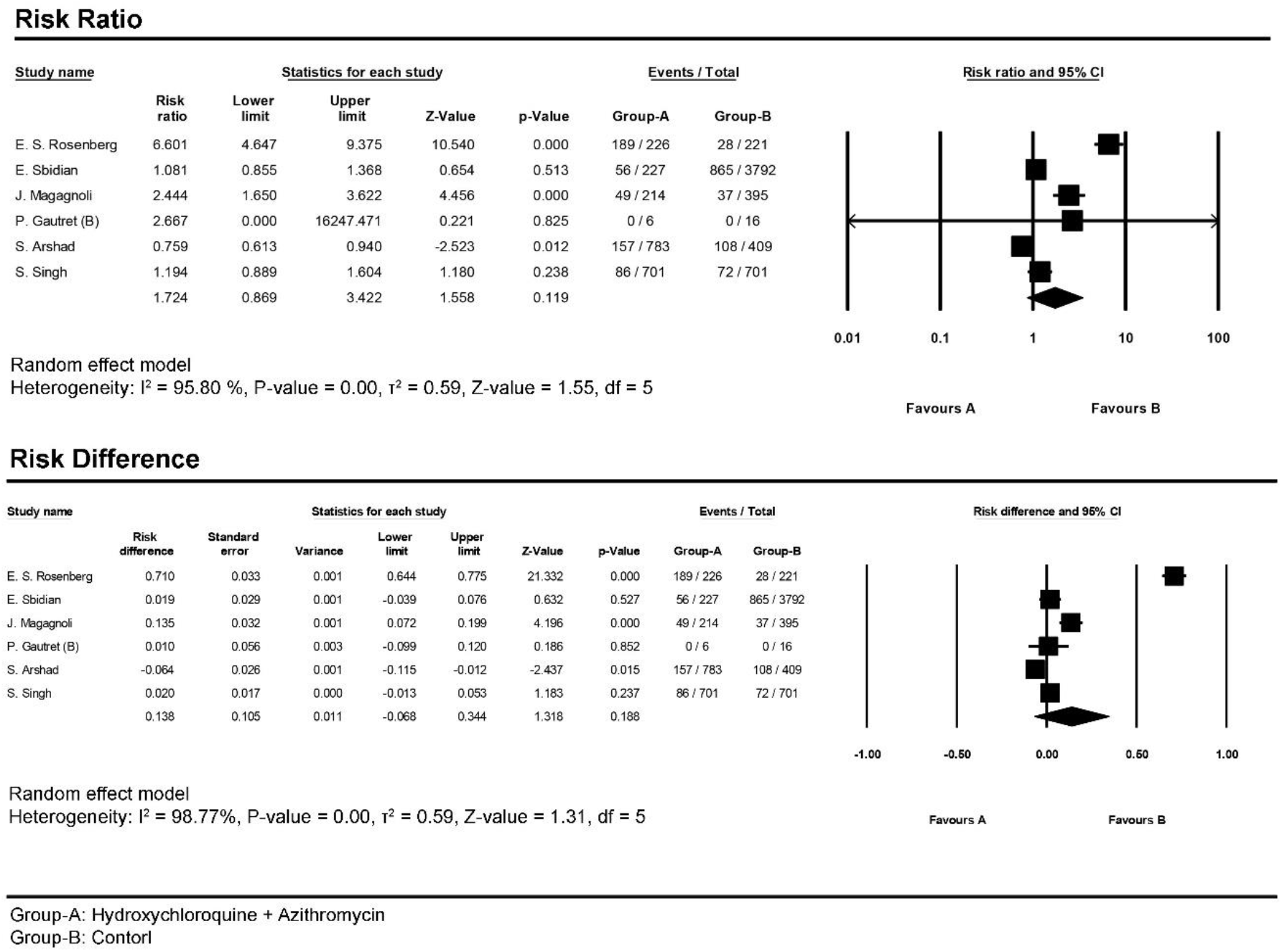
Forest plot for pooling risk ratios and risk differences regarding mortality rate in randomized and non-randomized studies

##### Overall mortality

For overall mortality, we considered the treatment arms of all comparative studies as observational studies and combined all types together as observational. The pooling mortality rate of investigations resulted in an overall mortality of 15.7% (95% CI, 13.1%-18.8%) for both HCQ and HCQ+AZM regimen (Fig. 9).

**Figure 9.**
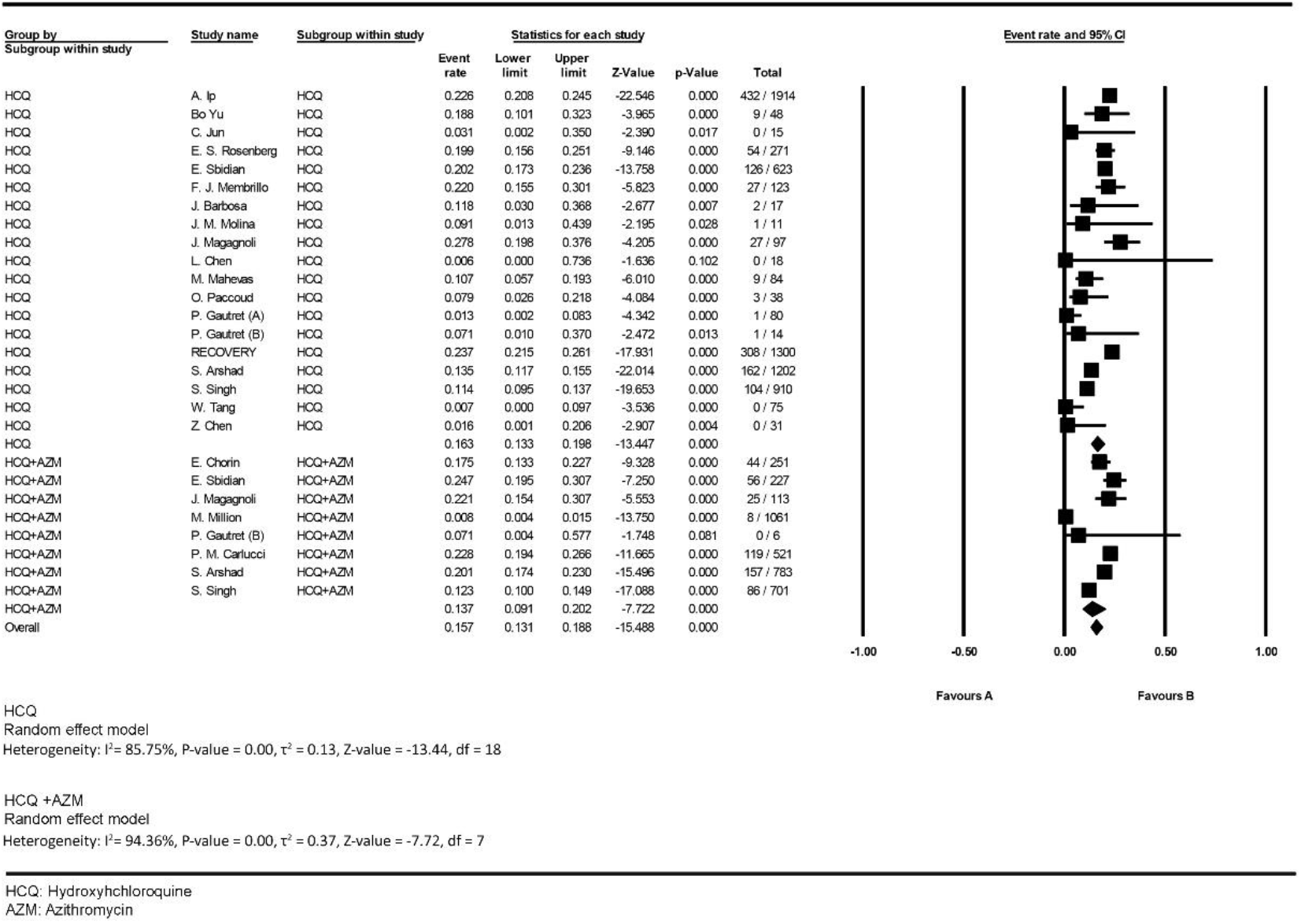
Forest plot for pooling mortality rates

##### Disease exacerbation

Meta-analysis of all comparative studies showed that the disease exacerbation was not significantly different between the HCQ group and the control group (RR: 1.23, 95% CI, 0.65-2.30; RD: 0.01, 95% CI, −0.08-0.11) (Fig. 10). Also, doing meta-analysis only on controlled randomized studies indicated no considerable disease exacerbation differences between two groups (RR: 0.59, 95% CI, 0.04-7.79; RD: −0.08, 95% CI, −0.32-0.14) (Fig. 11).

**Figure 10.**
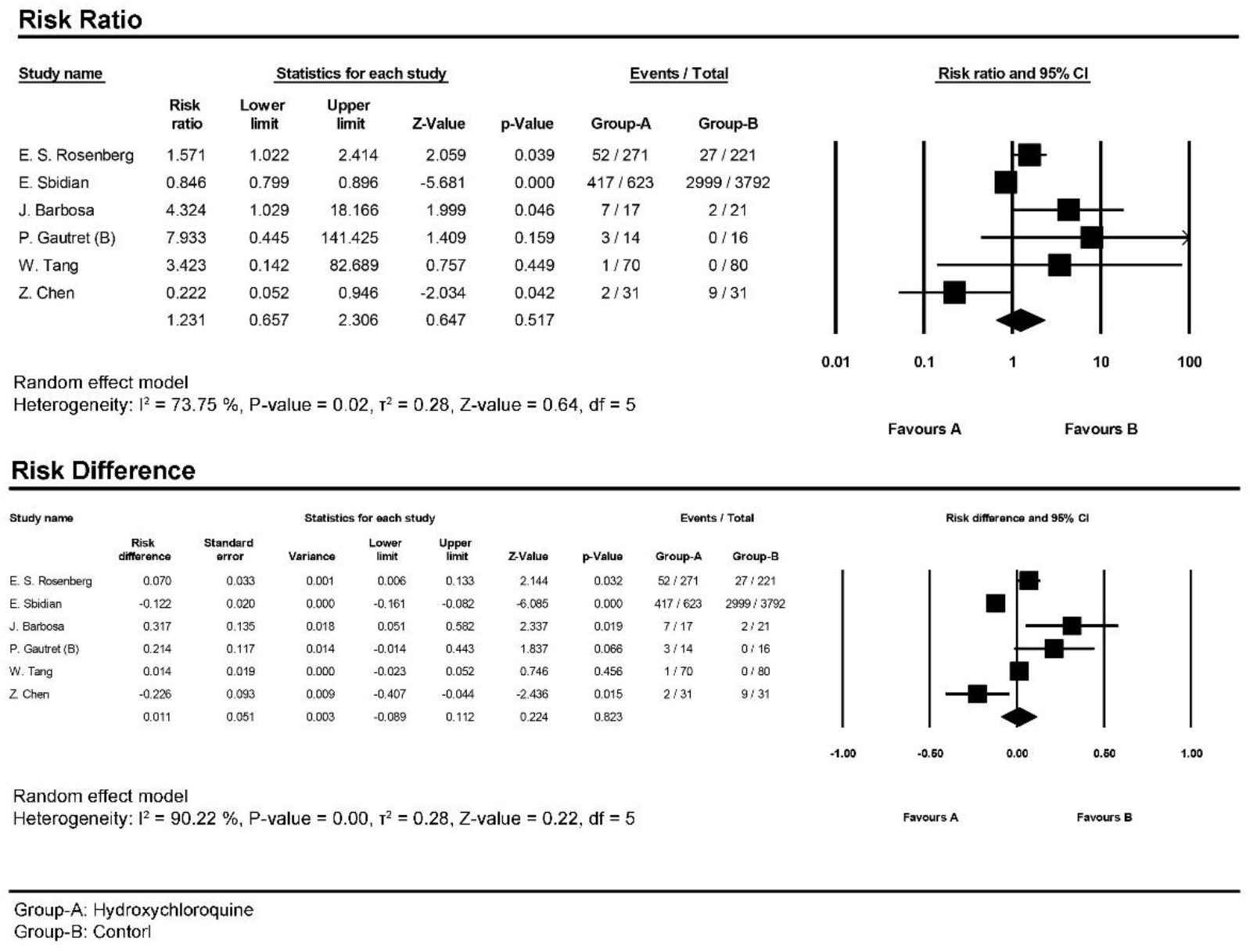
Forest plot for pooling risk ratios and risk differences regarding disease exacerbation in comparative randomized and non-randomized studies

**Figure 11.**
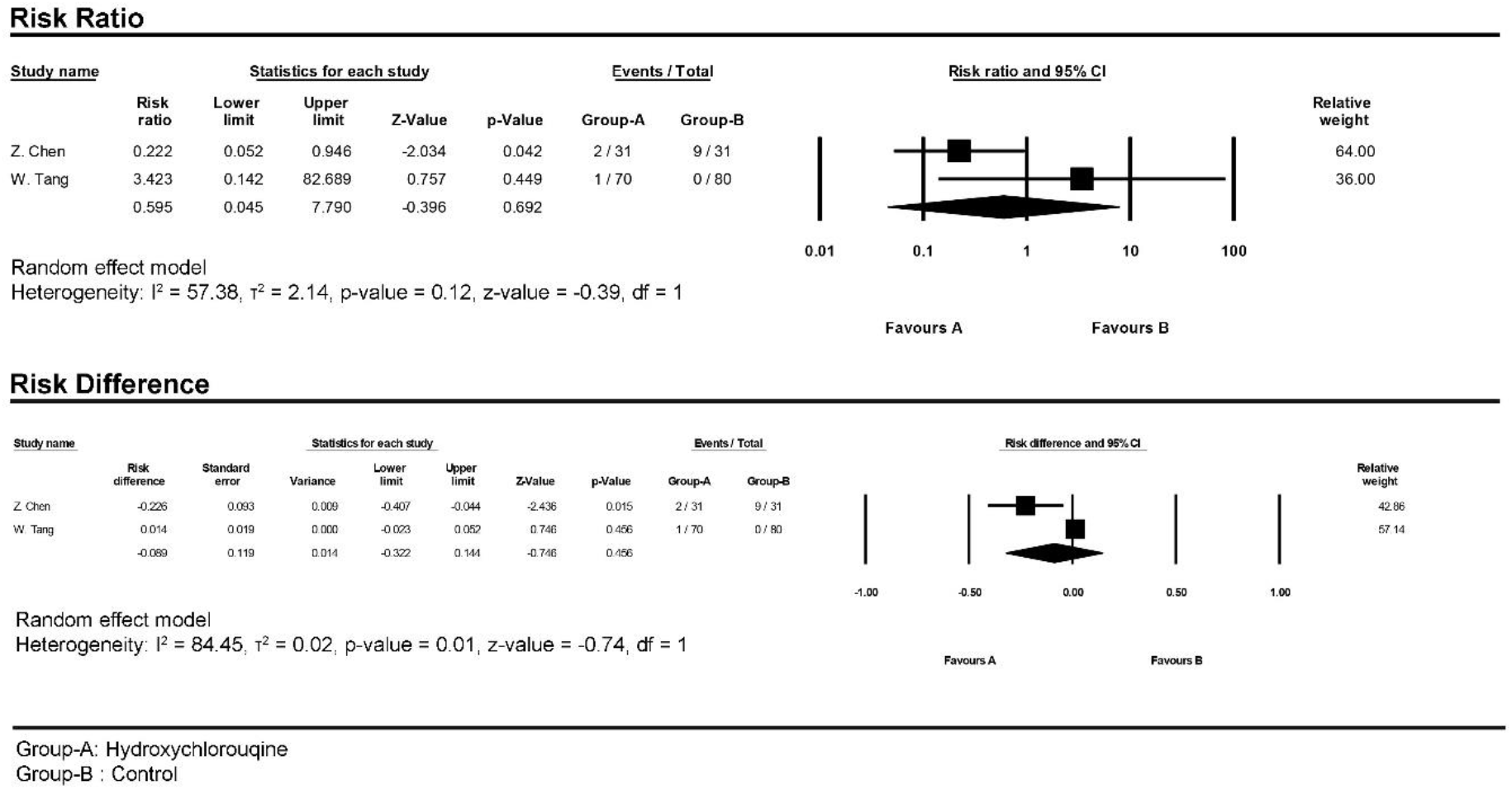
Forest plot for pooling risk ratios and risk differences regarding disease exacerbation in only controlled randomized studies

##### Intubation

Meta-analysis of comparative randomized and non-randomized studies indicated that there were no significant differences between the HCQ group and the control group about odds of intubation during treatment (OR: 2.11, 95% CI, 0.31-14.03) (Fig. 12).

**Figure 12.**
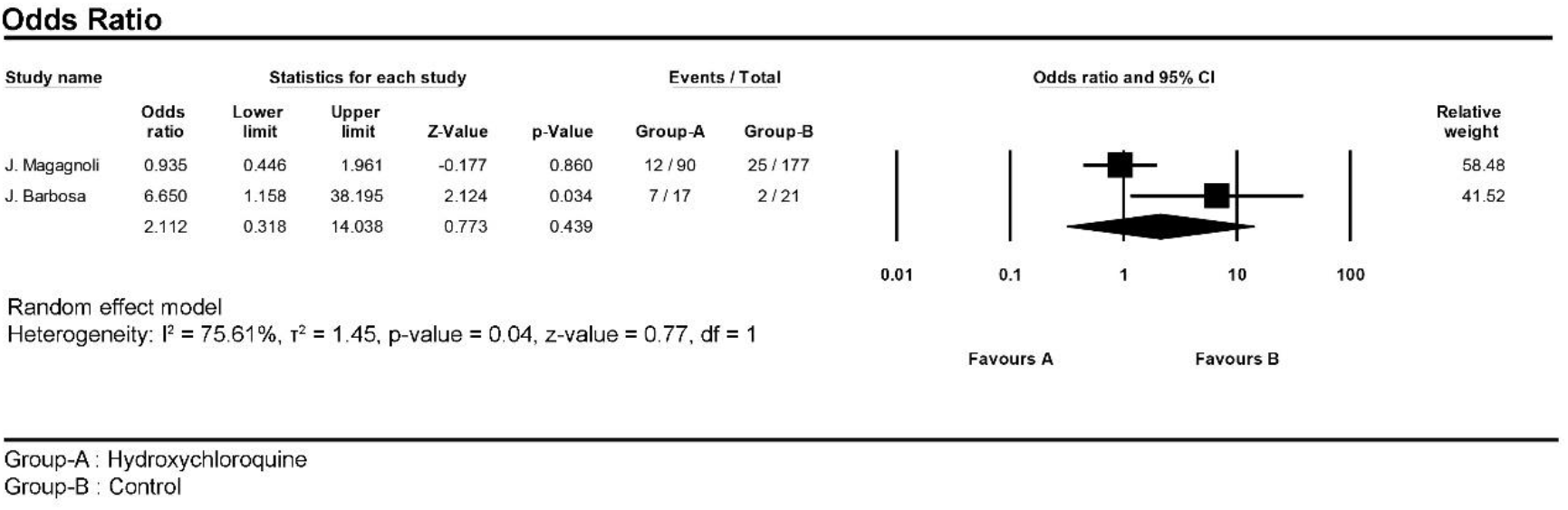
Forest plot for pooling odds ratios regarding intubation status

##### Adverse effects

The meta-analysis of comparative randomized and non-randomized studies showed that the odds of adverse effects occurrence in patients who received the HCQ regimen was approximately 3.5 times higher than the control group without HCQ regimen (OR: 3.40, 95% CI, 1.65-6.98) (Fig. 13). Likewise, doing meta-analysis only on controlled randomized studies indicated four times higher odds of experiencing adverse effects in patients who receive HCQ regimen in comparison to the control group (OR: 4.08, 95% CI, 1.84-9.04) (Fig. 14).

**Figure 13.**
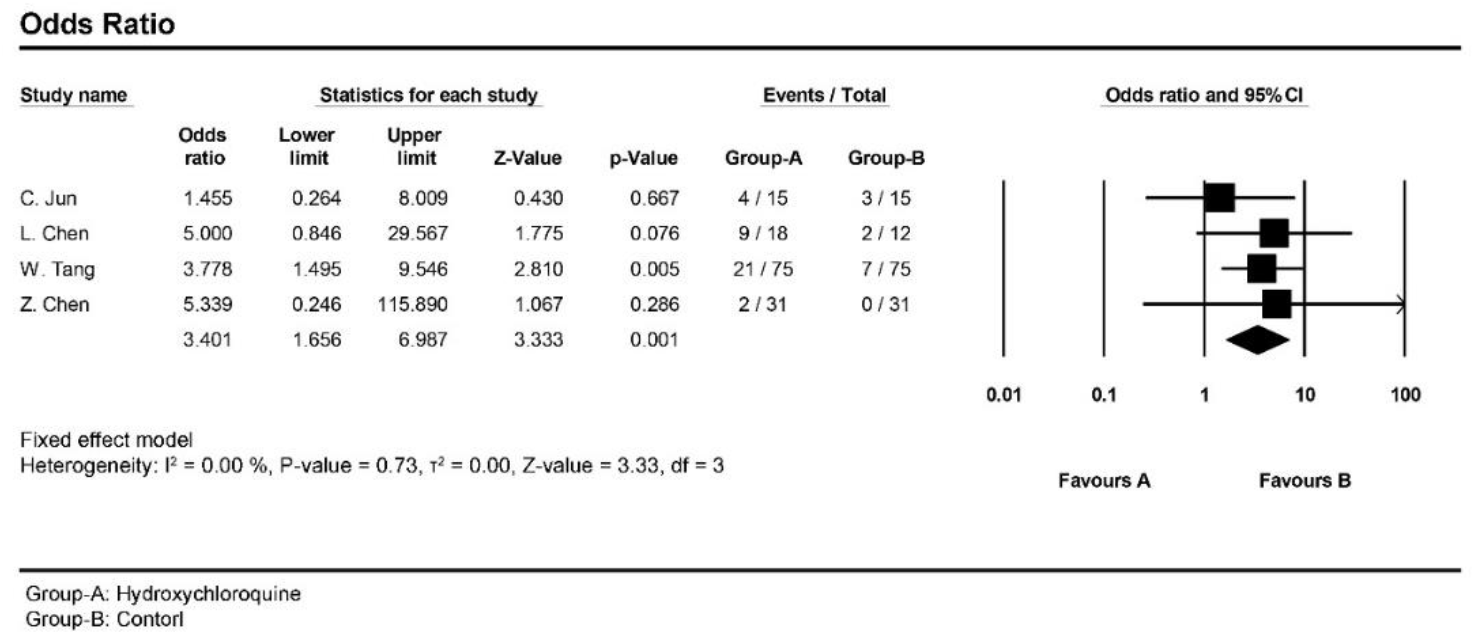
Forest plot for pooling odds ratios regarding adverse effects in comparative randomized and non-randomized

**Figure 14.**
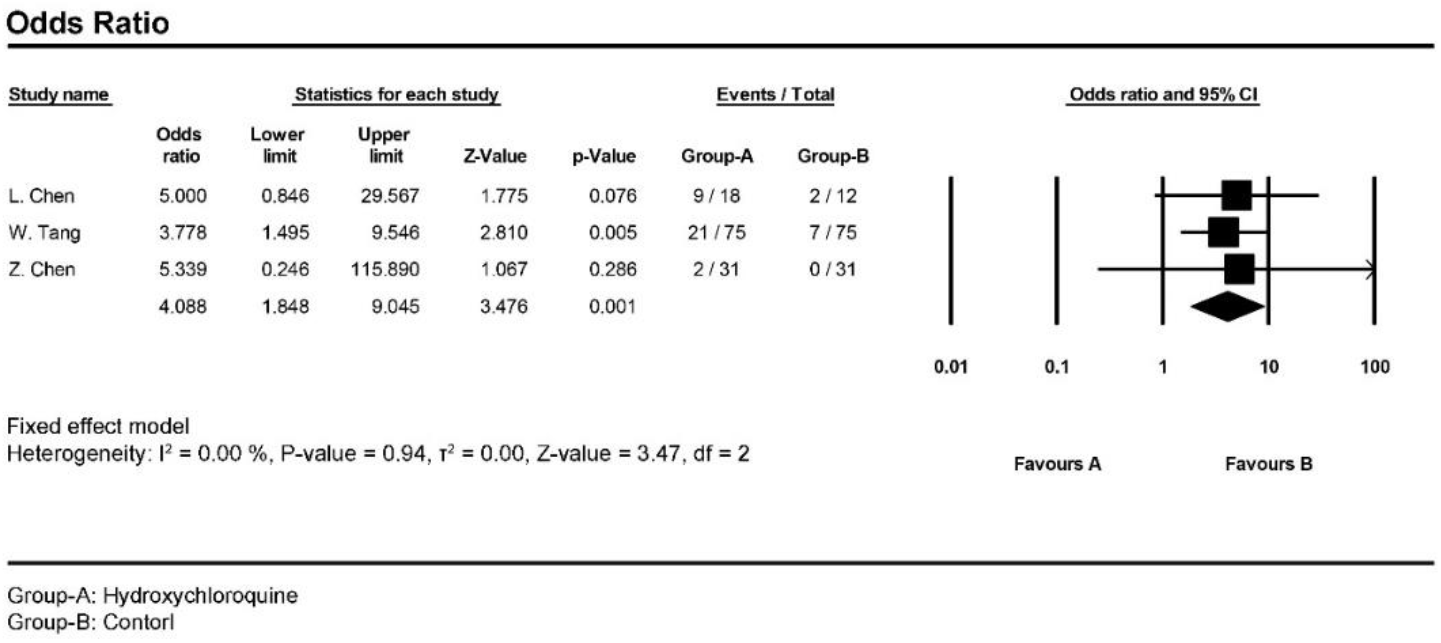
Forest plot for pooling odds ratios regarding adverse effects in only controlled randomized studies

##### Radiological improvement

A considerable Computed Tomography-Scan (CT-Scan) improvement was observed in the HCQ group with an odds ratio of 0.32 (95% CI, 0.11-0.98) (Fig. 15).

**Figure 15.**
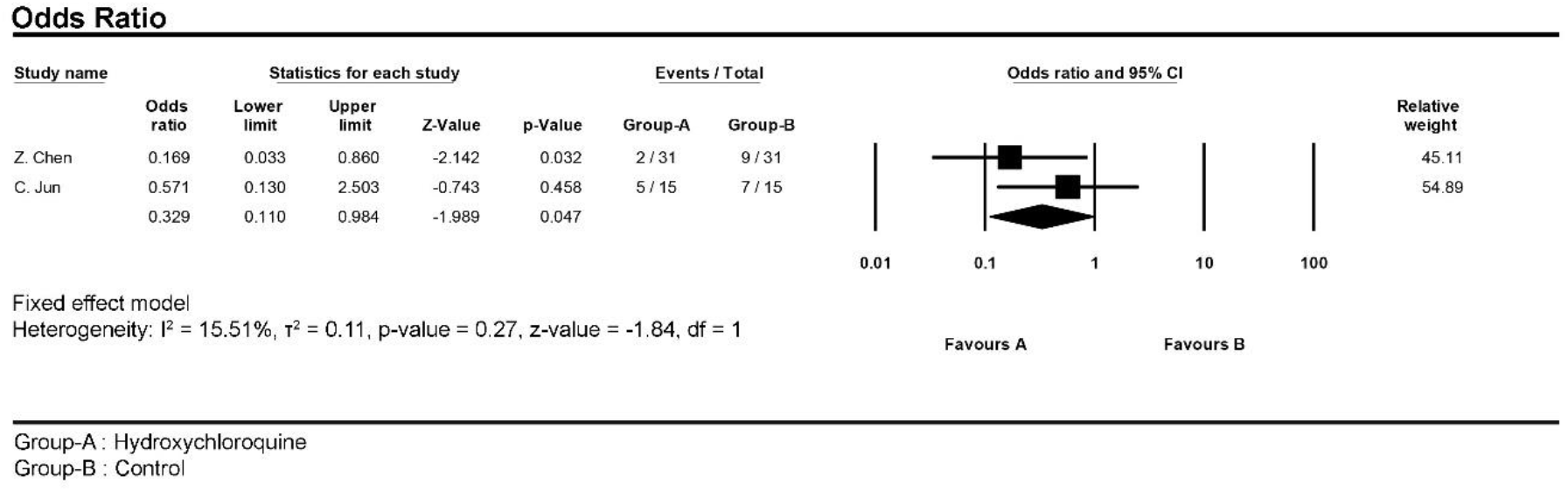
Forest plot for pooling odds ratios regarding radiological improvement

##### Meta-analysis of observational studies (Fig. 16)

We considered the treatment arms of the comparative studies as observational studies for this section. Hence, the meta-analysis of the other events rate showed the following results: 26.8% of patients suffered from known HCQ adverse effects (95% CI, 16.3%-40.7%); CT-Scan improvement has been observed in 16.5% of COVID-19 patients (95% CI, 2.8%-57.8%); 65.3% (95% CI, 56.7%-73.1%) of patients were discharged from the hospital or their nasopharyngeal culture resulted negative in RT-PCR evaluation, whereas 23.3% (95% CI, 8.9%-48.6%) of patients have been exacerbated, 7.1% (95% CI, 2.8%-17.0%) were admitted to the intensive care unit (ICU) and 23.8% (95% CI, 6.6%-57.9%) underwent intubation.

**Figure 16.**
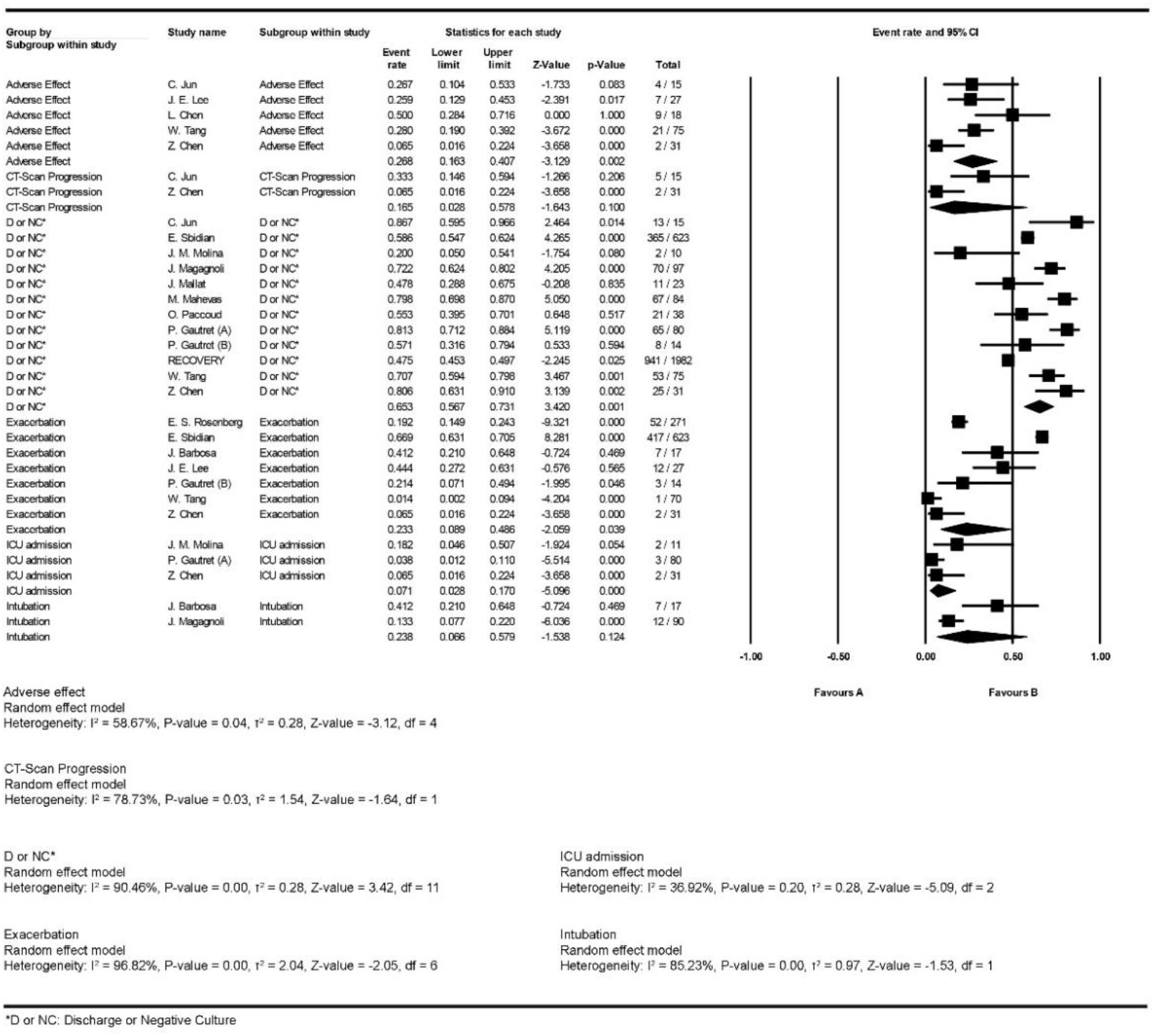
Forest plot for pooling events of observational studies

##### Prophylactic effects of hydroxychloroquine

Meta-analysis indicated no significant prophylactic effects for HCQ (OR: 0.40, 95% CI, 0.04-3.65) (Fig. 17).

**Figure 17.**
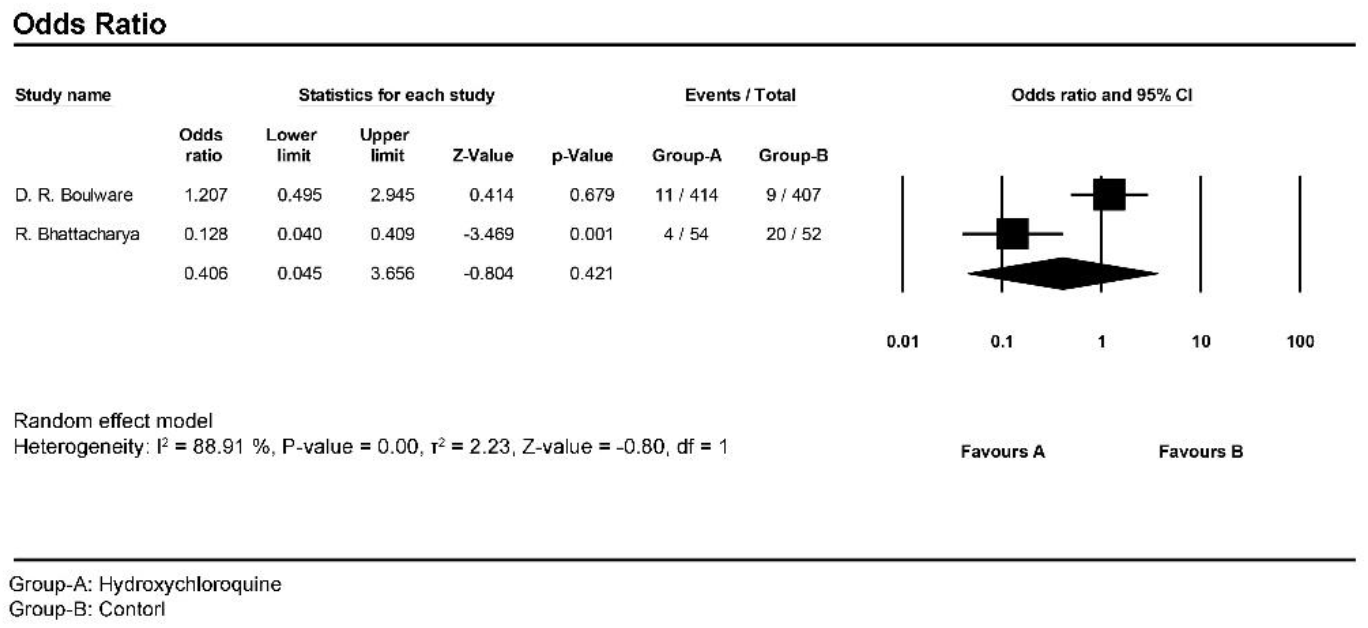
Prophylactic effects of hydroxychloroquine

## Discussion

More than five months after the closure of the Hubei region, there is still very little high-quality data on every treatment regimen, which raises questions about gaps in scientific works. In this context, one can only be surprised to see observational data without a control group covering a large number of patients when there is an essential need for randomized comparative data. In fact, Analysis of current data is difficult because at least 90% of spontaneous recovery is observed at the base. Thus, it takes well over 90% of recovery to consider that any treatment really brings something. The HCQ was no exception to this rule, with a result greater than 90%. On the other hand, in a few percents, the disease becomes serious with Acute Respiratory Distress Syndrome (ARDS) and multi-organ failure. This is where we would like to know what the treatments bring. Concerning all of the limitations and analyze difficulties, we have conducted this systematic review and meta-analysis with great caution and sensitivity in performing analyzes in order to try to overcome the current controversies regarding the effectiveness of HCQ in the treatment of COVID-19, at least at the base.

Considering the matter, recent investigations indicated that a high concentration of cytokines in the plasma called cytokine storm would be related to severe COVID-19 patients. In this situation, medications transposition is a critical need to find effective anti-inflammatory agents to decrease the cytokines and pro-inflammatory factors production ^45^. In this regard, HCQ has been known as an effective anti-inflammatory agent for a long time (since the 1950s), especially in autoimmune disorders ^46^. Besides, the outcome of a new experimental study conducted by Liu *et al*. has been mentioned in the title of their publication as follows: “*Hydroxychloroquine, a less toxic derivative of chloroquine, is effective in inhibiting SARS-CoV-2 infection in vitro*”. This also has been investigated and resulted same as the *in vitro* study of Yao *et al*. ^47^.

In addition, Pagliano *et al*. in their letter to the editor of *Clinical Infectious Diseases (CID)* journal, have been recommended the use of HCQ as pre/post-exposure prophylaxis against SARS-CoV-2 infection for health care staffs exposed to the virus in contaminated environments ^48^.

In contrast, Maurizio Guastalegname and Alfredo Vallone are claiming about the uselessness and even harmful effects of HCQ against COVID-19 in their letter to the editor in the above journal ^6^. They believe that, while the pathogenicity of the SARS-CoV-2 is still unknown, we should be cautious about the treatment decision, which has been proved through *in vitro* base studies in order to avoid dire paradoxical consequences like what has happened in treatment of *Chikungunya Virus* infection with chloroquine ^49^. Moreover, Molina *et al*. have followed 11 patients with HCQ + azithromycin regimen and concluded no clinical benefit and reasonable anti-viral activity ^19^. In addition, the pre-print of a Quasi-Randomized Comparative Study conducted in Detroit, Michigan, has been indicated not only any clinical benefits for HCQ but even increased need for urgent respiratory support (*p*=0.013) ^21^.

Also, H.J. Kim *et al*., in their opinion publication *for the COVID-19 Global Rheumatology Alliance* pointed at the shortage of HCQ following a sudden high demand after Gautret and colleagues’ publication on 20 March 2020 ^17^. They also referred to that HCQ is a crucial treatment choice for patients with systemic lupus erythematosus and rheumatoid arthritis disorders, who get into trouble in finding HCQ in this critical time ^50^. Authors recommend that scientific communities have to be very cautious and do not rush in the decision when there is no ample evidence for the subject, especially in such critical situations, which can lead to irreparable consequences. In fact, even if the efficacy of HCQ is confirmed, the world will be facing a new issue for both COVID-19 and rheumatic disorders patients: “*Shortage of Hydroxychloroquine*”.

More recently, the pre-printed study of Magagnoli *et al*. ^18^ on 368 United States veterans reported not only any clinical benefits for HCQ/HCQ+AZM regimen in COVID-19 patients but even association between higher mortality rate and HCQ group (hazard ratio: 2.61, 95% CI, 1.10-6.17; *P*=0.03). Also, the target trial emulation conducted by Mahévas *et al*. ^51^, did not support the effectiveness of the HCQ regimen, which has performed on 181 patients with SARS-CoV-2 hypoxic pneumonia.

Observational Study of Geleris *et al*. ^30^ in New York City on 1376 COVID-19 patients concluded that “*the study results should not be taken to rule out either benefit or harm of hydroxychloroquine treatment, given the observational design and the 95% confidence interval*”. However, due to that, investigators have combined the results of intubation and death together, we could not enter the results into meta-analysis. Another study conducted in New York State by Rosenberg *et al*. ^29^ on “*Association of Treatment With HCQ/AZM With In-Hospital Mortality in COVID-19 Patients*” has also similarly concluded no association for lower in-hospital mortality for both medications.

In addition, a multinational RECOVERY Collaborative Group with 4716 patients reported no associated for reductions in 28-day mortality in use of HCQ but increasing in length of hospital stay and risk of progressing to invasive mechanical ventilation or death.

In this case, we carried out the present systematic review in order to reach a clear result regarding taking or not-taking HCQ. In this study, the meta-analysis revealed no significant differences between HCQ arm who received HCQ regimen and standard treatment arm for both treatment effectiveness and mortality rate, which was the same for the HCQ+AZM regimen as well. For mortality, considering meta-regression analysis, age differences were significantly affected by obtained risk ratios. In addition, more cases in the HCQ group presented the improvement in CT-Scan results in comparison to the control group. Moreover, the considerably higher frequency of known HCQ adverse effects such as diarrhea, vomiting, blurred vision, rash, headache, etc. was observed in HCQ groups.

It is remarkable that Sarma *et al*. ^8^ conducted a meta-analysis on three studies and has been concluded the promising effects for HCQ clinical cure, which is inconsistent with our overall conclusion. Additionally, Million *et al*. ^7^ carried out “*a meta-analysis based on the first available reports*” released in *IHU Méditerranée Infection* (*COVID-IHU #10*), in which they have claimed a promising trend toward the chloroquine derivatives benefits against COVID-19 and suggested it to prescribe as a grade *I* recommendation. Going deeper into the content, the study suffers from some flaws such as ignoring heterogeneity as well as the pattern of dispersion in the results, combining different outcomes in an unusual way, using only odds ratios, when the risk ratios are the priority and preferred in such analysis, talking about four RCTs, whereas some are non-randomized, etc. Also, concerning the importance of mortality, Magagnoli *et al*. ^18^ findings, which go against the expected by authors, is ignored without any sensitivity analysis during the study.

As a matter of fact, “*statistics are only a tool for understanding and not an end in itself. If it is easy to say anything with statistics, it is also easy to say that you can make them say anything. However, using it well, with a strong consideration of the clinical sciences and an understanding of the possible sense of bias can go very far*”, Dr. Rauss says.

Our analysis did not indicate considerable effectiveness for the HCQ/HCQ+AZM regimen, which has been resulted in different studies, especially RECOVERY Collaborative Group multicenter, randomized, controlled trial. However, according to that, most of the studies are still observational; the possible influence of biases on the results in the analysis of such data is inevitable, but what is more important is to know in which direction the biases can play the role on the results. Thus, as we know, observational studies give a vision that overestimates the real result. Under these conditions, when we incorporate such data, and the result is not good, it means that we have a minimized vision of the result. Hence, the question arises whether the result might be even worse than what is observed?! If yes, then it would be understood that *“impact of this meta-analysis finings with strong consideration of the clinic and an understanding of the possible sense of bias is much greater than what is observed and presented”*; Dr. Rauss says.

In addition, based on recent evidence, the recommendation of Solidarity Trial’s International Steering Committee, WHO discontinued HCQ and lopinavir/ritonavir treatment arm for COVID-19 on 4 July 2020. However, it has been mentioned that “*This decision applies only to the conduct of the Solidarity trial in hospitalized patients and does not affect the possible evaluation in other studies of hydroxychloroquine or lopinavir/ritonavir in non-hospitalized patients or as pre- or post-exposure prophylaxis for COVID-19*” ^52^.

To our knowledge, this is the most updated systematic review that carried out a meta-analysis for investigating the role of HCQ with/without AZM in COVID-19 patients.

It is worth noting that the current meta-analysis includes the following limitations: 1) due to that most of the studies were non-randomized and results were not homogenous, selection bias was unavoidable; 2) various treatment plan regarding medication dosage and treatment duration; 3) insufficient moderator variables distribution by a group such as sex, underlying disease, etc.; and 4) inclusion of studies with small sample size, which leads to type *II* statistical errors. To overcome the limitations and bias, the results of the study should be confirmed by robustly randomized studies.

## Conclusion

This systematic review and meta-analysis showed no clinical benefits regarding HCQ treatment with/without azithromycin for COVID-19 patients. Although the mortality rate was not significantly different between cases and controls, the frequency of adverse effects was substantially higher in the HCQ regimen group. However, due to that, most of the studies were non-randomized, and results were not homogenous, selection bias was unavoidable, further large clinical trials following comprehensive meta-analysis should be taken into account in order to achieve more reliable findings. Also, it is worth mentioning that if this work does not allow to quantify a “value” of the HCQ, it allows at least to know what is not the HCQ and that it would be prudent not to continue investing in this direction. We hope our results could take a step toward this decision.

## Data Availability

The data that support the findings of this study are openly available in data bases mentioned in the search strategy.

## Acknowledgments

We would like to express our deepest appreciation to Dr. Alain Rauss, who extended a great amount of assistance in regards to the clinical and statistical contents of the study with his insightful advice and comments. Our gratitude should also go to the Student Research Committee of Mazandaran University of Medical Sciences for approving this student research proposal with the code 7586.

